# Epidemiological and Clinical Characteristics of Children with Coronavirus Disease 2019

**DOI:** 10.1101/2020.03.19.20027078

**Authors:** Qin Wu, Yuhan Xing, Lei Shi, Wenjie Li, Yang Gao, Silin Pan, Ying Wang, Wendi Wang, Quansheng Xing

## Abstract

**Background:** Severe acute respiratory syndrome coronavirus 2 (SARS-CoV-2) is a newly identified pathogen which mainly spreads by droplets. Most published studies focused on adult patients with coronavirus disease 2019 (COVID-19), but data concerning pediatric patients is limited. This study aimed to determine epidemiological characteristics and clinical features of pediatric patients with COVID-19.

**Methods:** We reviewed and analyzed data on pediatric patients with laboratory-confirmed COVID-19, including basic information, epidemiological history, clinical manifestations, laboratory and radiologic findings, treatment, outcome and follow-up results.

**Results:** From January 20th to February 27th of 2020, a total of 74 pediatric patients with COVID-19 were included in this study. Of the 68 cases whose epidemiological data were complete, 65 (65/68, 95.59%) cases were household contacts of adults whose symptoms developed earlier. Forty (59.46%) of the infected children were male, and the median age and body weight are 6 (0.10-15.08) years and 24 (4.20-87.00) kg, respectively. Except for one critically ill case, 20 (27.03%) patients did not show any symptoms of infection, 24 (32.43%) patients had acute upper respiratory tract infection and 29 (39.19%) patients were diagnosed with mild pneumonia. Cough (24/74, 32.43%) and fever (20, 27.03%) were the predominant symptoms of 44 (59.46%) symptomatic patients at onset of the illness. Abnormalities in leukocyte count were found in 23 (31.08%) children and 10 (13.51%) children presented with abnormal lymphocyte count. Of the 34 (45.95%) patients who had nucleic acid testing results for common respiratory pathogens, 19 (19 / 34, 51.35%) showed co-infection with other pathogens other than SARS-CoV-2.Ten (13.51%) children had RT-PCR analysis of SARS-CoV-2 for fecal specimens and 8 of them showed prolonged existence of SARS-CoV-2 RNA 11 (5-23) days after nasopharynx swabs turning negative. Abnormalities in chest imaging were observed in 37 (50.00%) patients and 28 (37.84%) of them only presented with nontypical changes.All children had good prognosis with a median hospitalization duration of 11 days and follow-up period of 16.5 days. During the follow-up period, all the patients remained in quarantine at designated site and home for two 14-day obervation periods and showed no clinical manifestation,but 3 of the 8 cases with prolonged fecal shedding of SARS-CoV-2 still showed positive result of feces test.

## Introduction

During the last three months, we have witnessed the fast-growing outbreak of coronavirus disease 2019 (COVID-19) swept through China and rapidly spread to all over the world. The etiological agent, severe acute respiratory syndrome coronavirus 2 (SARS-CoV-2), was identified as a novel pathogen with high infectiousness to general population and relatively high mortality rate. According to the lasted data released by the WHO on 17^th^ March 2020, 184,976 confirmed cases of COVID-19 were reported in 159 countries and regions globally, causing 7,529 deaths.^1^ Currently, the epidemic center has been shifted to Europe and WHO advocated that Europe should learn the lessons from China on infection control. Several COVID-19 studies from the Chinese researchers ^2-5^ have provided first-hand knowledge and valuable treatment experiences on the disease for other countries, but most of the studies were targeted at adult patients. Although pediatric cases of COVID-19 were constantly reported, the majority of published studies were case reports or studies with sample size of less than 50 pediatric cases. ^6-11^ To determine the spectrum of disease in children, we collected and analyzed epidemiological, clinical, laboratory, and radiologic data of 74 pediatric COVID-19 cases in two locations of northern and southern China. We hope our study would further the available knowledge regarding SARS-CoV-2 infection in children and provide an insight to treatment strategies and prophylactic control of the disease.

## Methods

### Data sources

From January 20^th^ to February 27^th^ of 2020, we retrospectively reviewed electronic medical records of 74 pediatric COVID-19 cases admitted in Qingdao Women’s and Children’s Hospital and Wuhan Children’s Hospital, including data recorded during hospitalization and follow-up period. Baseline information (gender, age, weight, time of onset, time of diagnosis by SARS-CoV-2 nucleic acid test, time of admission and discharge), epidemiological history, clinical manifestations, laboratory and radiologic findings, treatment, outcome and follow-up data were recorded with standardized data collection forms. This study was approved by the institutional review board of the ethics committee in our hospital (QFELL-KY-2020-11) and written informed consent was obtained from patients’ legal guardians prior to enrollment.

### Determination of exposure history

Detailed epidemiological data of all cases were collected and classified according to whether the case was household contacts of confirmed adult patients, the sequence of infection within the family, and whether the infected children transmitted the virus to others.

### Laboratory confirmation

Confirmation of COVID-19 was based on a positive result for real-time reverse transcription polymerase chain reaction (RT-PCR) testing of SARS-CoV-2 in nasopharynx swabs by hospital laboratory, and double-confirmed by local Centers for Disease Control and Prevention using the same RT-PCR protocol. Final decision on COVID-19 diagnosis was made according to WHO interim guidance. ^12^

### Diagnosis classification

According to the experts’ consensus statement on diagnosis, treatment, and prevention of 2019 novel coronavirus infection in children issued by group of respirology of Chinese Pediatric Society, ^13^ patients were classified into 5 types: asymptomatic infection, acute upper respiratory tract infection and mild pneumonia, severe pneumonia and critical cases (acute respiratory syndrome, SARS). The type of mild pneumonia was further sub-grouped into subclinical type and clinical type depending on whether the case had clinical manifestation.

### Statistical analysis

Categorical data were expressed as number (%) and continuous data were expressed as median with range or interquartile. Statistical analyses were performed using SAS software (SAS9.4; SAS Institute Inc, Cary, North Carolina, USA).

## Results

### Basic characteristics

None of the 74 infected children had comorbidities. Detailed data of baseline characteristics of the patients were listed in ***Table 1*** and ***Figure 1*** showed the timeline of disease progression including the following points in time: the date of admission, diagnosis, and discharge and the final date of follow-up.

**Table 1.** Basic and clinical characteristics of the study children infected with SARS-CoV-2,the data are expressed as n (%) or median (range).

| **Basic Characteristics** | No (percentage)/median (range) |
| --- | --- |
| Gender (F/M) | 30 (40.54%) /44 (59.46%) |
| Age, y | 6.00 (0.10-15.08) |
| $\leq 3$ months | 7 (9.46%) |
| $3 < \text{age} \leq 6$ months | 4 (5.41%) |
| $6 < \text{age} \leq 12$ months | 5 (6.76%) |
| $1 < \text{age} \leq 3$ years | 12 (16.22%) |
| $3 < \text{age} \leq 10$ years | 31 (41.89%) |
| Age $> 10$ years | 15 (20.27%) |
| Weight (kg) | 24.00 (4.20-87.00) |
| **Symptoms at onset** | 45 (60.81%) |
| Cough | 24 (32.43%) |
| Fever | 20 (27.03%) |
| Temperature, $^{\circ}\text{C}$, n=20 | 38.6 (37.6-40.10) |
| $\leq 38.0$ | 2 (10.00%) |
| $38.1-39.0$ | 10 (50.00%) |
| $> 39.0$ | 8 (40.00%) |
| Fatigue | 5 (6.76%) |
| Chest congestion | 4 (5.41%) |
| Anorexia | 3 (4.05%) |
| Diarrhea | 3 (4.05%) |
| Dyspnea | 2 (2.70%) |
| Headache | 2 (2.70%) |
| Expectoration | 2(2.70%) |
| Myalgia | 0 |
| Pharyngalgia | 0 |
| Dizziness | 0 |
| Myalgia | 0 |
| Clustering of the symptoms |  |
| 1 | 28 (37.84%) |
| ≥2 | 16 (21.62%) |
| **Laboratory findings** |  |
| White blood cell count, $\times 10^9/L$ | |
| 0-6 months | 7.43 (3.27-13.20) |
| 6 months-6 years | 8.22 (2.90-15.35) |
| ≥6 years | 6.80 (3.25-10.50) |
| Lymphocyte, $\times 10^9/L$ | |
| 0-6 months | 4.74 (1.91-8.10) |
| 6 months-6 years | 4.14 (0.80-9.03) |
| ≥6 years | 2.29 (0.86-4.54) |
| CRP, mg/L | 1.9(0.0-39.0) |
| abnormal | 13 (17.57) |
| PCT, ug/L | 0.10(0.03-0.75) |
| abnormal | 2 (2.70%) |
| ESR, mm/h | 18 (12-25) |
| abnormal | 5 (5/14, 35.71%) |
| Co-infection | 19 (19/34, 51.35%) |
| mycoplasma | 16 (84.21%) |
| RSV | 3 (15.79%) |
| EBV | 3 (15.79%) |
| CMV | 3 (15.79%) |
| influenza A&B | 1 (5.26%) |

CRP: C-reactive protein, PCT: procalcitonin, ESR: erythrocyte sedimentation rate, RSV: respiratory syncytial virus, EBV: Epstein-Barr virus, CMV: cytomegalovirus.

**Figure 1.**
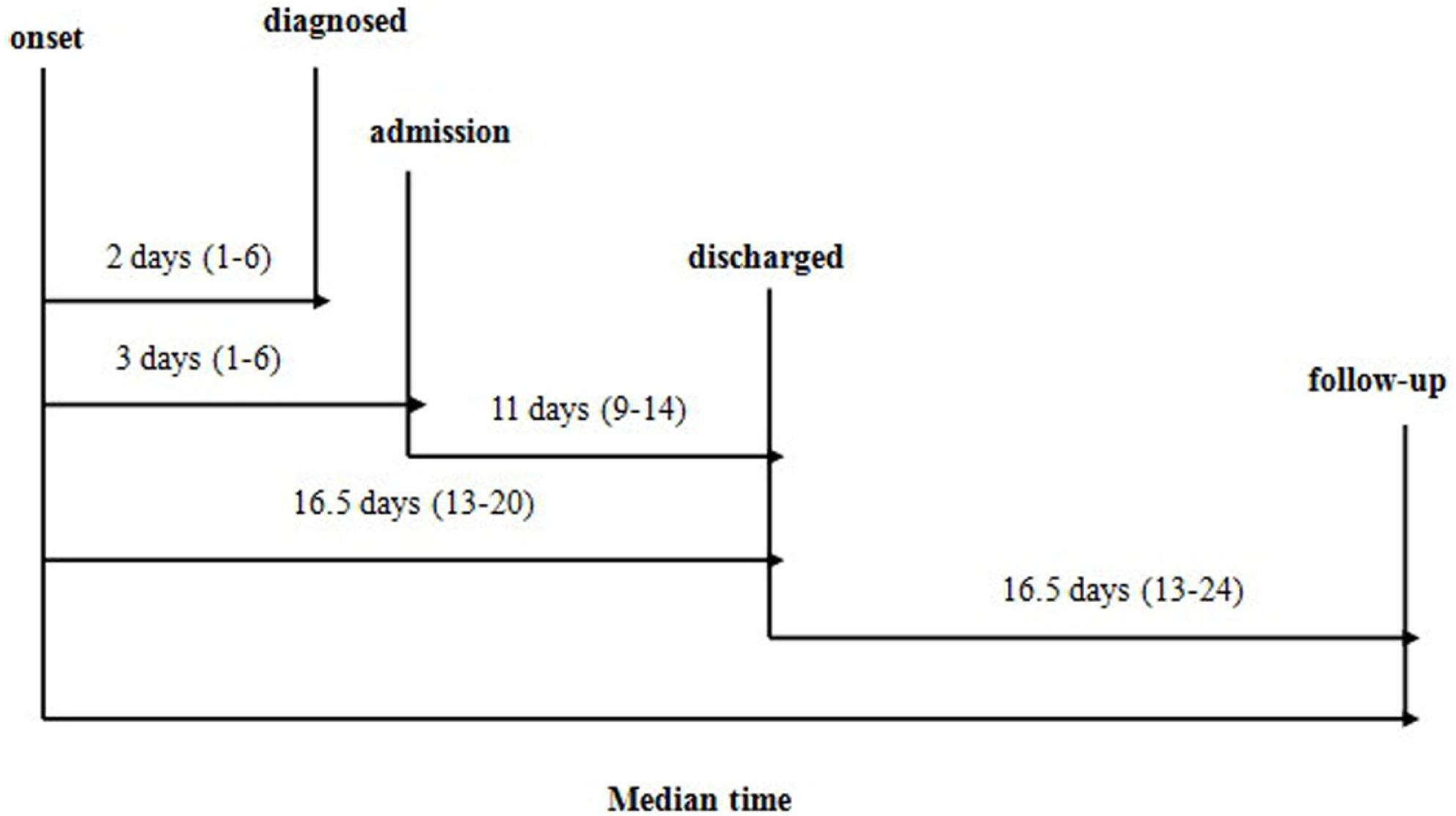
Timeline of COVID-19 cases after onset of illness. The interval time was expressed as median with interquartile.

### Epidemiological characteristics

Complete information of exposure history was collected from 68 of the 74 patients (91.89%). Except for 3 sporadic cases, 65 (65/68, 95.59%) cases were household contacts of adults whose symptoms developed earlier and the last confirmed case within the family, including 18 (27.69%) being the second infected family member, 23 (35.38%) being the third, 14 (21.54%) being the fourth, 9 (13.85%) being the fifth and 1 (1.54%) being the sixth. There has been no evidence showing the virus was transmitted from children to others.

### Clinical features

#### Diagnosis and classification

There was only one severe pneumonia case among the 74 infected children, and the rest consisted of 20 cases of asymptomatic infection, 24 acute upper respiratory tract infection cases and 29 mild pneumonia cases, accounting for 27.03%, 32.43% and 39.19% respectively. Thirty (40.54%) children did not have clinical symptoms and were identified by SARS-CoV-2 nucleic acid screening test, and 20 of them were finally confirmed as asymptomatic carriers whereas 10 children were classified as subclinical mild pneumonia patients.

#### Clinical manifestations

Cough (32.43%) and fever (27.03%) were the most common symptom at the onset of disease. Other symptoms like fatigue, chest congestion, anorexia, diarrhea, dyspnea, headache and expectoration were seldom; myalgia, pharyngalgia, dizziness and myalgia was rather rare. Detailed data are listed in ***table 1***. Except fever, all the positive signs were related to respiratory system, including rhonchi and crackles in 16 cases (21.62%). No children showed obvious positive signs of nervous system and digestive system. The most severe case was a 13-year-old boy with body-weight of 85 kg who presented with high fever and cough at onset of the disease, and his fever lasted for 3 days with a highest body temperature of 39.8 LJ. During hospitalization, he suffered from dyspnea with low proximal oxygen saturation of 92% on room air and had bilateral diffuse breath sounds of crackles which could be heard for 5 days.

#### Laboratory findings (table 1)

There were 23 cases (31.08%) with abnormal leukocyte count, being increased in 19 cases (25.68%) with a highest level of 15.35 × 10^9^/L and reduced in 4 cases (5.41%) with a bottom level of 2.90 × 10^9^/L. Abnormal lymphocyte count was found in 10 cases (13.51%), among whom 6 (8.11%) had increased number of lymphocytes (highest count 9.03 × 10^9^/L) and 4 (5.41%) had reduced number of lymphocytes (lowest count 0.80 × 10^9^/L). C-reactive protein (CRP) was increased in 13 cases (17.57%) with a highest serum level of 39.0mg/L. Elevation of procalcitonin (PCT) was observed in 2 cases (2.70%) with a highest level of 0.75ug/L. Erythrocyte sedimentation rate (ESR) was measured for 14 (18.92%) patients and 5 (5/14, 35.71%, 12-25 mm/h) of them had accelerated results. Of 74 cases, 34 (45.95%) were screened for common respiratory pathogens and 19 of them (19/34, 51.35%) had co-infection, including 11 cases co-infected with mycoplasma pneumoniae (MP), 2 with MP and respiratory syncytial virus (RSV), 2 with MP and Epstein-Barr virus (EBV), 2 with cytomegalovirus (CMV), 1 with CMV and EBV, and 1 with MP, influenza A&B and RSV. Other routine laboratory testing showed no obvious abnormality. Ten (13.51%) children had RT-PCR analysis for SARS-CoV-2 in fecal specimens and viral RNA remained positive in stools of 8 convalescent patients after respiratory specimens showing negative. The time for SARS-CoV-2 RNA in fecal specimens turning negative after negative conversion in nasopharynx swabs ranged from 5 to 23 days, with a median of 11days.

#### Radiologic findings

Four (5.41%) children only had chest computed tomography (CT) examination once which was done on admission and did not show any abnormalities. The other 70 (94.59%) children included in this study had chest CT examination both at the time of admission and discharge. Radiological changes were found in 37 (50.00%) patients, with 8 (8/37, 21.62%) cases in left lung, 13 (13/37, 35.14%) cases in right lung and 16 (43.24%) cases bilaterally. Of the 37 cases with abnormal chest CT findings, 30 cases (81.08%) had clinical symptoms, and 7 (7/37, 18.92%) showed no symptoms during the whole course of illness. There were only 9 cases (9/74, 12.16%) with typical changes of SARS-CoV-2 infection in chest CT imaging, including patches of ground glass opacity which were mainly distributed near the pleura (***Figure. 2***). Changes in CT imaging of the other 28 cases was nonspecific for SARS-CoV-2 infection (***Figure. 3***). No large area of “white lung”, pleural effusion and pneumothorax was found among the patients. Chest CT imaging of the most severe case only showed abnormalities of common viral pneumonia (***Figure. 4***).

**Figure 2.**
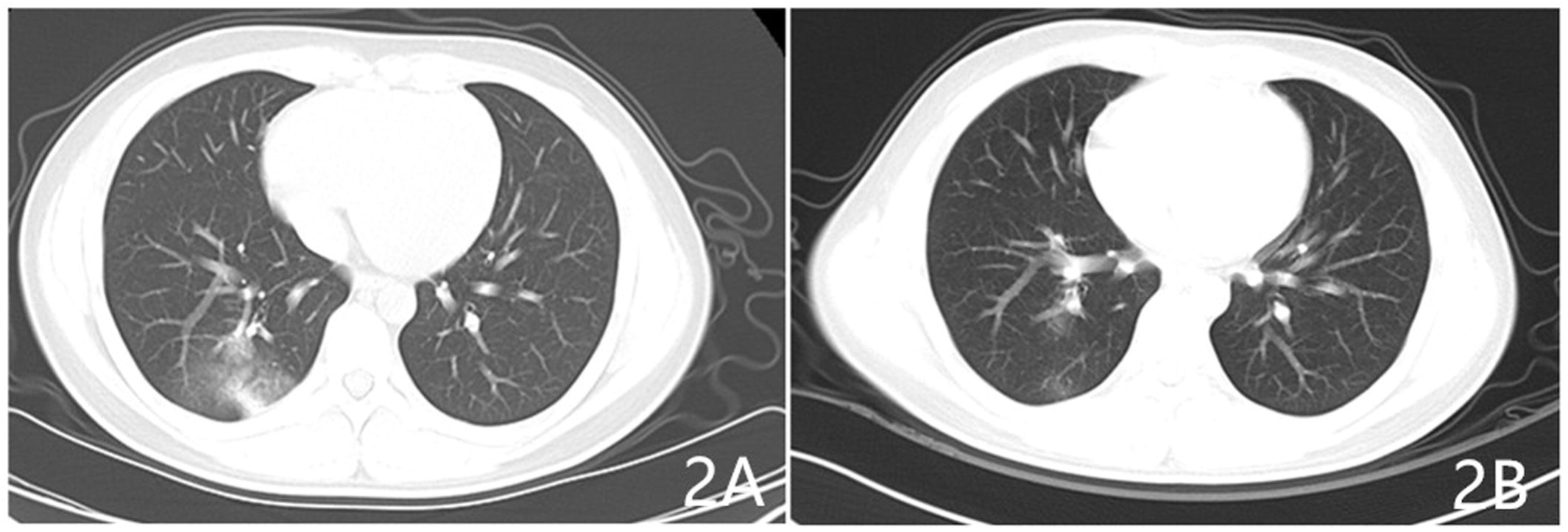
Chest CT images from a 2-year-old boy showed ground glass opacity near the pleura of the lower lobe of right lung on day 2 after symptom onset (2A) and the lesion almost was absorbed after 12 day’s treatment (2B).

**Figure 3.**
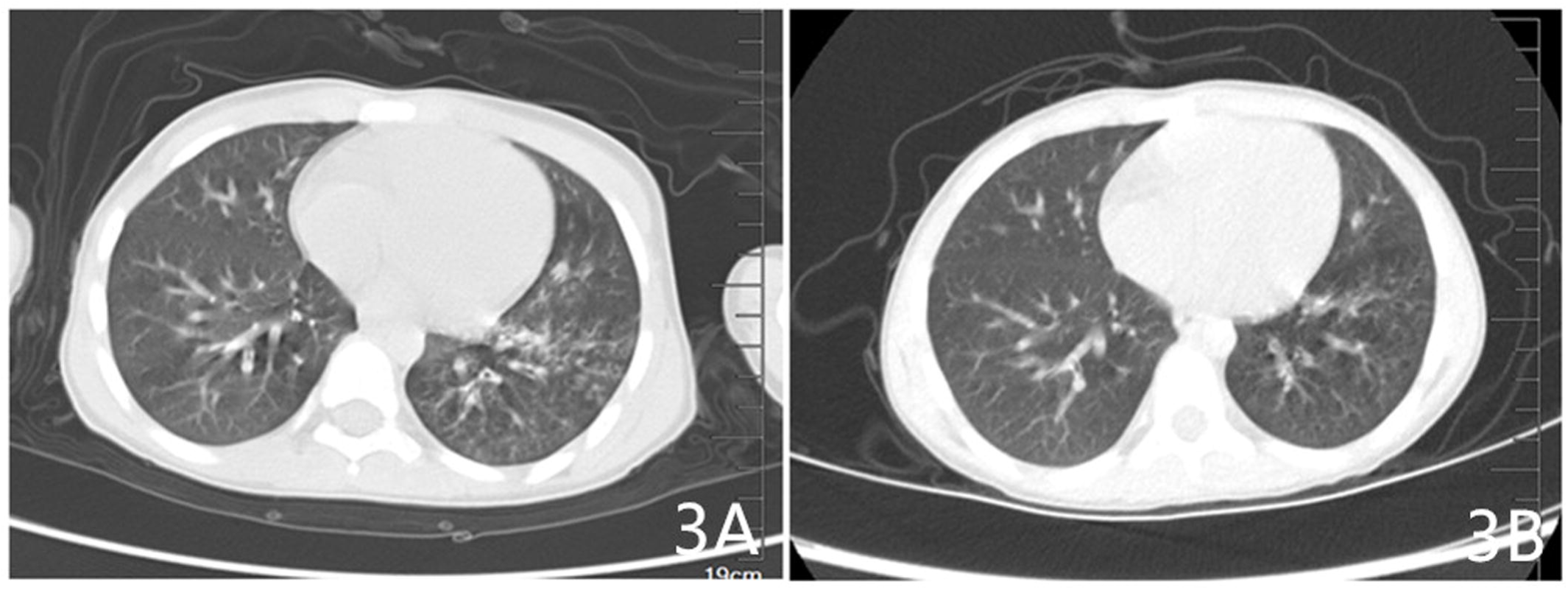
Chest CT images from a 7-year-old girl showed bilateral nonspecific multiple patches along the bronchovascular bundle on day 5 after symptom onset (3A) and the lesion was absorbed after 8 day’s treatment (3B).

**Figure 4.**
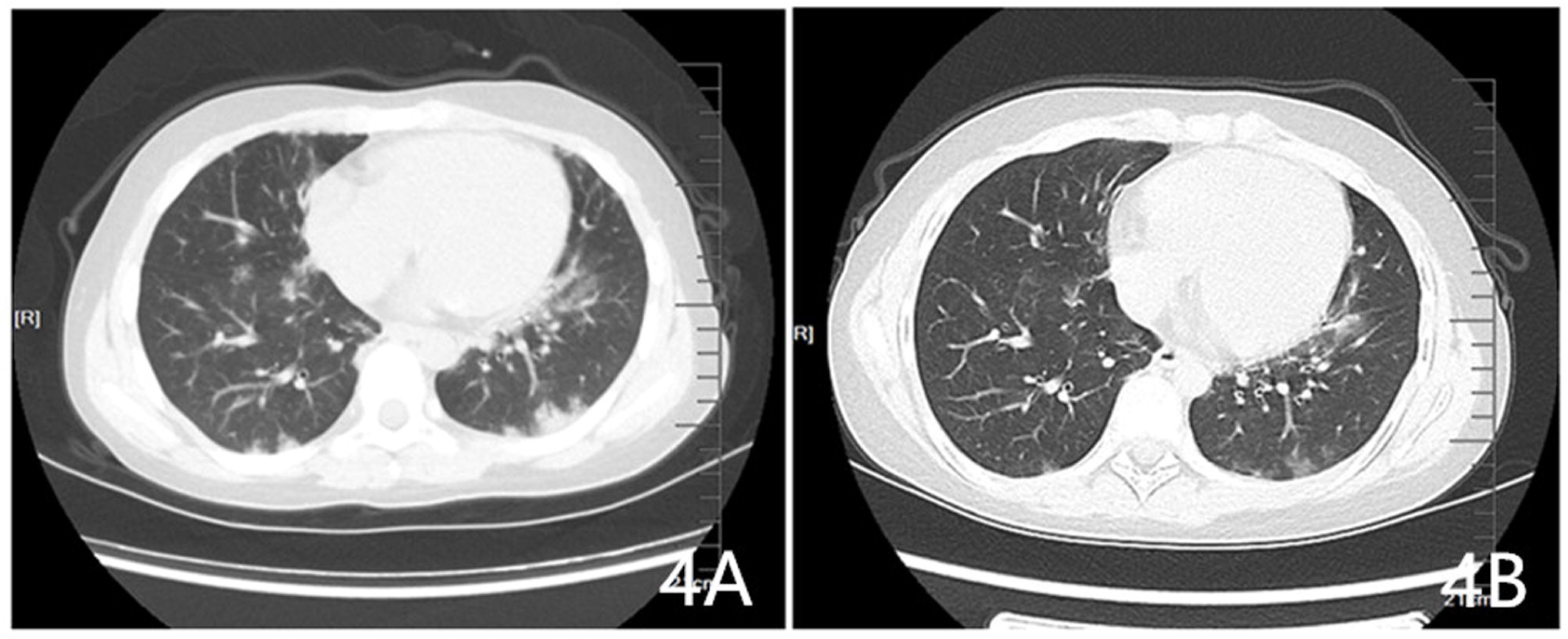
Chest CT images from the severe patient of a 13-year-old boy showed bilateral nonspecific multi-patchy consolidation opacity near the pleura on day 6 after symptom onset (4A) and the lesion was absorbed after 11 day’s treatment (4B).

#### Treatment and outcomes

All the 74 cases were treated according to the experts’ consensus statement on diagnosis, treatment, and prevention of 2019 novel coronavirus infection in children issued by group of respirology of Chinese Pediatric Society, ^13^ including interferon inhalation, administration of antiviral drugs and Traditional Chinese Medicine (TCM). Fifteen patients with confirmed MP infection were treated with azithromycin orally or intravenously, and the other 12 children received empirical antibiotic therapy. Only the 13-year-old severe case was given systematic corticosteroids for 5 days and gamma globulin for 3 days. No patient required mechanical ventilation except one severe child who received noninvasive ventilation for 5 days. All the 74 patients were discharged based on the tentative fifth or sixth edition for Diagnosis and Treatment of COVID-19 (tentative fifth or sixth edition) issued by the National Health Commission of the People’s Republic of China ^14, 15^ with good prognosis.

#### Follow-up results

After being discharged from the hospital, all patients were remained in quarantine at designated sites for a 14-day medical observation and then sent for home confinement for another two weeks. At the time of writing, the median follow-up period of patients in this study was 16.5 (10-42) days. No convalescent patients discharged from hospital showed clinical manifestation during the study period, but 3 of the 8 cases with prolonged fecal shedding of SARS-CoV-2 were still positive for RT-PCR analysis till the last day of follow-up.

## Discussion

The newly issued Report of the WHO-China Joint Mission on Coronavirus Disease 2019 ^16^ pointed out the attack rate in individuals under 18 years of age was only 2.4% and no death was reported in this age group. According to the data released by China Centers for Disease Control and Prevention (China CDC), ^17^ 416 cases were reported among children 10 years old and younger with no death case, which accounted for 0.9% of total COVID-19 cases. Data from local health authorities showed the attack rate among children ranged from 4.9% to 7.6%. ^18, 19^ Evidence so far implied that children are less severely affected by COVID-19, which resembles that of SARS emerged 17 years ago. ^20^ Familial cluster is one of the common features of COVID-19 in children. ^7, 9, 10, 21^ Among the 74 pediatric cases included in this study, 68 had complete exposure history recorded and 65 (95.59%) were household contacts of adults whose symptoms developed earlier. There has been no evidence showing the virus was transmitted from children to others. However, the relatively low attack rate of COVID-19 in children might be explained by the stringent implementation of home confinement and nation-wide school closure as required by the Chinese governments. During the outbreak, public activities were discouraged, and children spent most of their days at home with strengthened protection from caregivers.

This study included clinical, laboratory, treatment, and outcome data of 74 pediatric cases of COVID-19 from two children’s hospitals within and beyond epidemic center (from southern China and northern China, respectively). All patients were discharged from the hospital after recovery and were followed up for 14 days. Clinical presentations of infected children were distinct from that of adult patients. Except for one critically ill patient, 20 (27.03%) cases were asymptomatic carriers of SARS-CoV-2, 53 (71.62%) were mild to moderate cases with various manifestations. Among adult patients, fever (83.0%-98.6%) and cough (59.4%-82.0%) were the most common and predominant symptoms. ^2-5^ Whereas fever and mild cough only accounted for 27.03% and 32.43%, respectively, of symptoms at disease onset and during hospitalization in our pediatric patients, a proportion much lower than that of adult patients. Additionally, fatigue, headache, nausea, and gastrointestinal symptoms were not common among infected children. Nearly half of the children were not admitted to hospital for symptoms; they were found SARS-CoV-2 positive for RT-PCR analysis during quarantine after family members had been diagnosed with COVID-19.

Laboratory findings of pediatric patients were also different from adult patients. Previous studies have shown leukopenia, lymphopenia, and increased serum levels of CRP in majority of adult patients. ^2-5^ Only one third of the 74 pediatric cases in our study had abnormal leukocyte and/or lymphocyte counts, among whom 19 (25.68%) had increased and 4 (5.41%) had decreased number of leukocytes; 6 (8.11%) had elevated and 4 (5.41%) had reduced lymphocytes. Moreover, magnitude of these abnormalities in leukocytes and lymphocytes was relatively small without typical changes. Given that children under 6 years of age have higher lymphocyte counts than adults, further consideration would be warranted for evaluation of these indexes. There was no pattern for the changes in other infection-related variables such as CRP, PCR and ESR.

Typical radiologic changes in adult COVID-19 patients include multifocal areas of ground glass shadows and infiltration in both lungs, and dynamic changes could be observed as the disease progresses. Abnormalities in lung imaging could be served as one of the criteria for assessment of disease severity and prognosis according to the Trial Guideline for Diagnosis and Treatment of COVID-19 (tentative fifth & sixth & seventh edition) issued by the National Health Commission of the People’s Republic of China. ^14, 15, 22^ However, pediatric cases of COVID-19 lacked the typical changes in chest imaging. ^11^ Among the 74 cases in this study, only 9 (12.16%) of the 37 cases who showed abnormalities in CT imaging developed into ground glass opacity; whereas the rest 28 cases only showed atypical changes for bronchopneumonia and common viral pneumonia. Nearly half of the cases did not show any Radiologic changes during course of the disease.

The exact reason for why children are less severely affected is still unclear. One possible explanation is related to children’s immature immune system. Their adaptive immunity is less developed to mount an inflammatory response during immune dysregulation phase induced by viral infection. ^23^ Relatively stronger humoral responses in children may also contribute to this phenomenon. Innate immunity is able to react more rapidly in response to pathogen invasion than adaptive immunity. Moreover, children generally have fewer comorbidities, making them more resilient to SARS-CoV-2 infection. Most pediatric patients had relatively mild disease with good prognosis, which could also be observed in children infected with SARS-CoV and other respiratory viruses. Compared with pediatric patients, mortality rate of seasonal flu in adults is nearly 10 times higher. ^24^ Several studies have shown that children with SARS only presented with fever, cough and nasal congestion, and seldom developed into the third phase of SARS (characterized by acute respiratory distress syndrome). Accordingly, it is difficult to distinguish children with SARS from infection with other common respiratory viruses. ^25-27^ Non-specific clinical, Radiologic and laboratory characteristics of children with SARS-CoV-2 infection making them hard to be recognized, raising the possibility of misdetection of asymptomatic carriers. A comprehensive evaluation of epidemiological exposure and nucleic acid testing results would be warranted to guide clinical decision making.

More attention should be drawn to children with COVID-19 who also have co-infection with other common respiratory pathogens. Among the 74 pediatric patients included in this study, 19 (51.35%) of the 34 children who had testing results for common respiratory pathogens had co-infection; 8 (42.11%) children had two or more pathogens other than SARS-CoV-2 detected. This finding was consistent with our previous observation and also in line with studies of others. ^28, 29^ It can be seen that co-infection is common in children with COVID-19, indicating SARS-CoV-2 screening is necessary to identify COVID-19 cases, especially during the peak season for colds, influenza and other respiratory ailments.

With the accumulating experience of COVID-19 management in China, an increasing number of studies have reported positive RT-PCR results for fecal SARS-CoV-2 detection or isolation of viable virus from excretions. ^30, 31^ Xiao and colleagues demonstrated that viral RNA could exist in stools of COVID-19 patients for 12 days or longer. ^30^ Previously we found that RT-PCR testing results for SARS-CoV-2 remained positive in feces of three pediatric patients for approximately 4 weeks, a duration much longer than that of respiratory specimens (about 2 weeks).^32^ Here we simultaneously conducted nucleic acid testing in nasopharynx swabs and fecal specimens for 10 of the 74 pediatric patients and all of them had positive results in both samples. Eight children had fecal SARS-CoV-2 positive for RT-PCR analysis after negative conversion of viral RNA in respiratory specimens. At the time of writing, three children still had fecal RNA detectable and the longest lagged behind for 23 days. SARS-CoV-2 may present in gastrointestinal tract for a longer time than respiratory system, which appears to be more common among pediatric patients. However, there is limited data on comparison with adult patients. Detection of viral RNA in feces of convalescent patients does not necessarily mean that the virus are replication-competent nor infectious enough to be transmitted. There is still a possibility of fecal-oral transmission. The emerging disease brings new challenges to preparedness response and prophylactic control, in particular, massive efforts should be made at all levels to minimize spread of the virus among children after reopening of kindergartens and schools.

Personalized management should be considered for pediatric patients with COVID-19, more precisely, to symptomatic and asymptomatic carriers of SARS-CoV-2 who come from families with a cluster of infection. Treatment plan should also be tailored for children. ^33^ Anti-symptomatic therapies including administration of antipyretics and anti-viral agents, inhalation of interferon, and enhanced nutrition support were given to children with fever and pneumonia. Only one critically ill case was given low dose of systemic corticosteroid in a short period of time. Over half of the pediatric patients were merely under close surveillance and quarantine and none of them had worsening of symptoms. The most ‘severe’ symptom during their hospitalization and medical observation was the one at onset of the illness. More investigations are needed to determine the standardized treatment for pediatric patients with typical COVID-19 symptoms and to evaluate the efficacy of traditional Chinese medicine in children.

### Limitations

The rapidly evolving COVID-19 outbreak and the lack of specific containment measures in the early stage caused panic in the community and hospitals. In such cases, we failed to collect the complete epidemiological information from 6 patients due. Only blood routine, biochemical and infection-related biomarkers were analyzed in this study due to the different standards for laboratory testing between the two hospitals. We were unable to measure the viral loads, or to detect presence of SARS-CoV-2 in nasopharynx swabs and fecal specimens for all of our patients. We only screened common respiratory pathogens for children who were admitted to hospitals in the later stage. But we believe these 74 cases with complete medical records during hospitalization and follow-up period represented a certain proportion of pediatric patients of COVID-19 in China.

## Data Availability

I had full access to all the data in the study and took responsibility for the integrity of and the accuracy of the data.

## Contributors

YW, W-DW and Q-SX had full access to all the data in the study and took responsibility for the integrity of and the accuracy of the data. YW, W-DW, Q-SX, QW, LS and Q-SX designed the study and conceptualized the paper. QW and LS collected the epidemiological, clinical, laboratory and radiological data. W-JL, YG and S-LP contributed to laboratory testing. LS and Q-SX summarized the data. QW, LS, Y-HX and Q-SX wrote the initial draft of the manuscript. All authors provided critical feedback and approved the final version.

## Acknowledgements

This work was funded by The National Natural Science Foundation of China (NSFC) [Grant number 81770315], Distinguished Taishan Scholars (2019), Qingdao Outstanding Young Health Professional Development Fund (2020) and Qingdao People’s Livelihood Science and Technology Program(17-3-3-6-nsh). We thank all patients and their families involved in the study. We thank all healthcare workers involved in the diagnosis and treatment of patients.

